# Spatially resolved transcriptomic profiling for glomerular and tubulointerstitial gene expression in C3 glomerulopathy

**DOI:** 10.1101/2023.06.30.23292064

**Authors:** Jung Hun Koh, Minji Kang, Sehoon Park, Jeong Min Cho, Semin Cho, Yaerim Kim, Soojin Lee, Hajeong Lee, Kwon-Wook Joo, Yon Su Kim, Kyung Chul Moon, Hyun Je Kim, Dong Ki Kim

## Abstract

**Introduction:** C3 glomerulopathy (C3G) is a rare but clinically significant glomerulopathy. However, little is known about its transcriptomic profile. We investigated the substructure-specific gene expression profile of C3G using the recently introduced spatial transcriptomics technology.

**Methods:** We performed spatial transcriptomic profiling using GeoMx Digital Spatial Profiler with formalin-fixed paraffin-embedded kidney biopsy specimens of three C3G cases and seven controls from donor kidney biopsy. Additionally, 41 samples of other glomerulonephritis, including focal segmental glomerulosclerosis, membranous nephropathy, and minimal change disease, were included as disease controls. Gene expression levels were compared by DESeq2 method to identify differentially expressed genes (DEGs). We performed gene ontology (GO) annotation through the ToppGene suite and mapped interactions among the DEGs using the STRING database.

**Results:** We identified 229 and 157 highly expressed DEGs in the glomeruli of C3G compared to those of donor and disease controls, respectively, with consistently highest fold changes in POSTN, COL1A2, and IFI44L. Protease binding, structural molecule activity, and extracellular matrix structural constituent were among the top enriched GO terms in the glomeruli of C3G, with consistent features seen in the network analysis. In contrast, no significant GO enrichment was found among the 563 and 347 lowly expressed DEGs in the glomeruli of C3G compared to the controls. The tubulointerstitial transcriptomic profiles of C3G were similar to those of the controls.

**Conclusion:** In the glomerulus of C3G, genes related to the extracellular matrix and interferon activity were the most upregulated. Significant disease-specific transcriptomic alterations in C3G provide potential insights into the pathophysiology.

## Introduction

C3 glomerulopathy (C3G) is a primary glomerulonephritis that results from dysregulation of the complement system.^1^ Although it remains a rare disorder with incidence estimated at a few cases per million, 30 to 50 percent of the cases progress to end-stage kidney disease within a decade of diagnosis, and up to 60% of patients experience recurrence after kidney transplantation.^2,3^ Current medical management is limited to supportive care for proteinuria along with selective use of immunosuppressive agents and plasmapheresis with mixed reports on their efficacy.^4^ While complement inhibitors have been proposed as potential disease-specific therapies, recent studies of the C5 inhibitor eculizumab in C3G have not shown consistent efficacy.^5,6^

The recent recognition of C3G as a distinct histopathological entity has allowed more precise characterizations of the underlying pathophysiology. Studies based on familial cases as well as animal models showed that constitutive activation of the alternative pathway is key to the development of the disease, mediated by factors including mutations in complement regulator genes and autoantibodies to enzymes in the complement cascade.^1,7^ At the same time, components of the glomerular microenvironment, which include the glomerular glycocalyx and epithelial cells, interact with complement regulators and appear to play a crucial role in local regulation of the complement pathway.^1,8^ Given the structural complexity of the glomerular microenvironment, however, how the relevant molecular pathways integrate and manifest as C3G remains to be elucidated.

One evolving approach in addressing such questions in kidney diseases has been to employ spatial transcriptomics.^9^ Spatial transcriptomics technologies enable mapping of gene expression profiles to specific histological structures, which may be as small as individual glomeruli.^9,10^ Spatial transcriptomics profiling of the kidney in C3G may reveal patterns of gene expression that indicate the most relevant molecular pathways and networks in the pathophysiology of the disease.

In the present study, we employed the spatial transcriptomics analysis framework to analyze gene expression patterns in the glomeruli and tubules of C3G. We hypothesized that C3G would have distinct glom- or tubule-specific transcriptomic changes in comparison to healthy donor kidneys and to other common glomerular diseases.

## Methods

### Ethical considerations

This study was approved by the Institutional Review Board of Seoul National University Hospital (IRB No. 2208-137-1353). Patient-matched formalin-fixed paraffin-embedded (FFPE) samples of C3G and other glomerular diseases used as diseases controls were provided from the study hospital, as the hospital stores remnant tissue blocks after establishment of the diagnosis. Informed consent was waived by the Institutional Review Board as the study retrospectively used remnant tissues and health records. The study was conducted in accordance with the Declaration of Helsinki.

### Sample selection and collection

Study samples were selected from kidney biopsies performed at Seoul National University Hospital between 2009 and 2021 and stored as FFPE blocks. Samples with pathologic diagnosis of minimal change disease (MCD), membranous nephropathy (MN), focal segmental glomerulosclerosis (FSGS), and diabetic nephropathy (DN) were included as disease controls. Healthy control specimens were obtained from time-zero allograft biopsies performed during living donor kidney transplantation. To enhance detection of changes specific to each glomerular disease, the inclusion criteria were set as follows: (1) at least 18 years of age, (2) inclusion of 10 or more glomeruli per section, (3) estimated glomerular filtration rate (eGFR) of 30 mL/min/1.73 m^2^ or greater, (4) free of histopathologic features of DN in non-DN cases. The inclusion criteria for donor controls were further specified to have preserved kidney function with eGFR of 80 mL/min/1.73 m^2^ or greater and be free of any preexisting glomerular diseases under pathologic review.

### Slide preparation and processing

Kidney tissues were mounted on the Leica BOND Plus slides with thickness of 5 μm. The sections on the slides were baked at 60°C for 30 minutes, deparaffinized with CitriSolv and gradually rehydrated with ethanol and 1X phosphate-buffered saline (PBS). To unmask the antigen epitopes, the slides were dipped in 1X Tris-EDTA (pH 9.0) and incubated for 15 minutes, followed by RNA target exposure with a 1 μg/mL proteinase K solution at 37°C for 15 minutes and wash in 1X PBS for 5 minutes. For *in situ* hybridization, the slides were then incubated overnight at 37°C with the GeoMx Whole Transcriptome Atlas, which consists of RNA detection probes conjugated with unique ultraviolet-photocleavable oligonucleotide barcodes targeting 18,677 target genes. The slides were subsequently washed then stained with the following morphology markers for 1 hour at room temperature: SYTO 13 (Nanostring, 121300303), Pan-cytokeratin (Novus, NBP2-33200 AF594), and alpha-smooth muscle actin (Abcam, ab202296).

The slides were loaded into the GeoMx Digital Spatial Profiler (DSP) instrument, and representative glomerular and tubulointerstitial substructures were selected as regions of interest (ROIs) by a kidney pathologist. The oligonucleotide barcodes for the target genes within each ROI were photocleaved and collected into a DSP collection plate. The oligonucleotides were amplified through polymerase chain reaction (PCR) with primer pairs and i5 and i7 dual-indexing sequences. The PCR products were pooled and purified with AMPure XP Reagent (A63880) to obtain the sequencing library. Quality of the library was assessed with the Agilent 4150 TapeStation system, and they were subsequently sequenced on an Illumina NovaSeq 6000 with 27 × 27 paired-end reads.

### Bioinformatics and statistical analysis

Raw sequencing reads in FASTQ format were converted into digital count conversion files using the GeoMx NGS Pipeline software (v2.0). The DSP Data Analysis Suite (v2.4) was used for data analysis and quality control. Genes that were not expressed in over 50 percent of the included samples were excluded from the downstream analysis.

After filtering, unnormalized read counts were used to draw relative log expression plots to identify technical outliers. To identify differentially expressed genes (DEGs), we used the DESeq2 package in R (version 3.6.2).^11^ We compared the differences between the C3G and donor controls, and between C3G and other glomerular disease controls. Genes with a false-discovery rate of less than 0.05 by the Benjamini–Hochberg correction were considered significant DEGs for analysis. The upregulated and downregulated significant DEGs were analyzed using the ToppGene Suite mainly for functional enrichment analysis based on Gene Ontology (GO) annotations.^12^ To investigate the protein-protein interaction networks among the DEGs, we used the STRING database.^13^ The DEGs were mapped based on interaction scores, and DEGs with at least one significant interaction with other DEGs were presented in the graphs.

## Results

### Clinical characteristics

Three cases of C3G were included in the study, as were 7 donor controls and 41 other glomerulonephritis samples of MCD, MN, FSGS, and DN as disease controls. The clinical characteristics and pathological features of the C3G cases are described in **Table 1**. Of note, all three cases were male with the mean age of 61.3 years, and all patients presented with proteinuria but with variable degree of azotemia, edema, and hematuria. Pathological diagnosis in all three cases was consistent with C3 glomerulonephritis, and while the proportion of sclerotic glomeruli was variable, there were sufficient non-sclerotic glomeruli for spatial transcriptomics analysis. Five years after the initial diagnosis, renal function was stable in one patient, while eGFR declined by 47% to stage G4 chronic kidney disease in another patient. The third patient was referred out after one year of stable renal function.

**Table 1.**
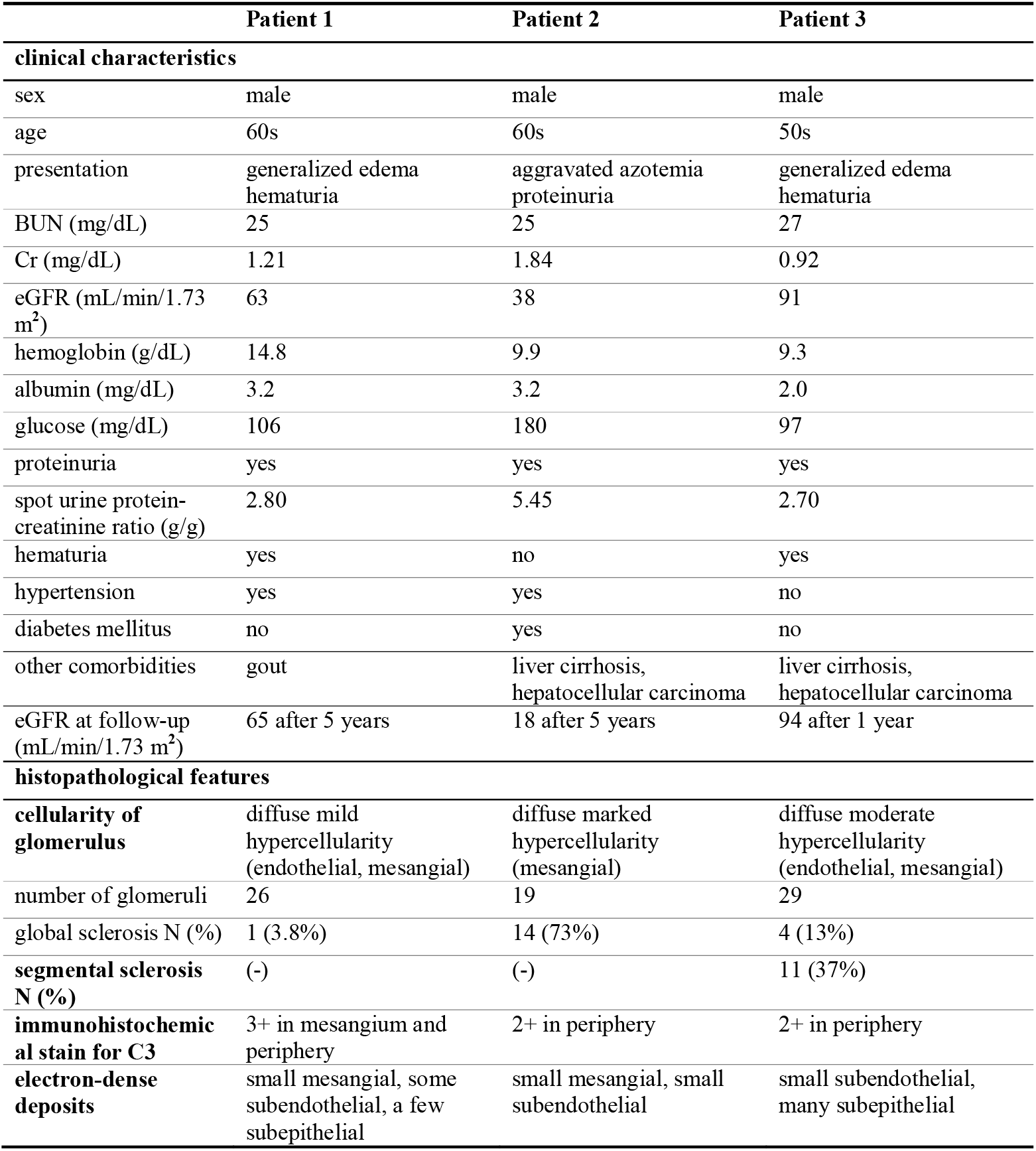
Clinical characteristics and histopathological features of C3 glomerulonephritis cases included in the study. (BUN: blood urea nitrogen; Cr: creatinine; eGFR: estimated glomerular filtration rate; N: number)

|  | Patient 1 | Patient 2 | Patient 3 |
| --- | --- | --- | --- |
| **clinical characteristics** |  |  |  |
| sex | male | male | male |
| age | 60s | 60s | 50s |
| presentation | generalized edema<br>hematuria | aggravated azotemia<br>proteinuria | generalized edema<br>hematuria |
| BUN (mg/dL) | 25 | 25 | 27 |
| Cr (mg/dL) | 1.21 | 1.84 | 0.92 |
| eGFR (mL/min/1.73 m<sup>2</sup>) | 63 | 38 | 91 |
| hemoglobin (g/dL) | 14.8 | 9.9 | 9.3 |
| albumin (mg/dL) | 3.2 | 3.2 | 2.0 |
| glucose (mg/dL) | 106 | 180 | 97 |
| proteinuria | yes | yes | yes |
| spot urine protein-creatinine ratio (g/g) | 2.80 | 5.45 | 2.70 |
| hematuria | yes | no | yes |
| hypertension | yes | yes | no |
| diabetes mellitus | no | yes | no |
| other comorbidities | gout | liver cirrhosis,<br>hepatocellular carcinoma | liver cirrhosis,<br>hepatocellular carcinoma |
| eGFR at follow-up (mL/min/1.73 m<sup>2</sup>) | 65 after 5 years | 18 after 5 years | 94 after 1 year |
| **histopathological features** |  |  |  |
| **cellularity of glomerulus** | diffuse mild hypercellularity (endothelial, mesangial) | diffuse marked hypercellularity (mesangial) | diffuse moderate hypercellularity (endothelial, mesangial) |
| number of glomeruli | 26 | 19 | 29 |
| global sclerosis N (%) | 1 (3.8%) | 14 (73%) | 4 (13%) |
| segmental sclerosis N (%) | (-) | (-) | 11 (37%) |
| **immunohistochemical stain for C3** | 3+ in mesangium and periphery | 2+ in periphery | 2+ in periphery |
| **electron-dense deposits** | small mesangial, some subendothelial, a few subepithelial | small mesangial, small subendothelial | small subendothelial, many subepithelial |

Clinical characteristics of the healthy donor controls and other glomerular disease controls are summarized in **Table 2**. All healthy donor controls had preserved eGFR, while 33 of 41 (80%) of disease control cases had eGFR of 45 mL/min/1.73m^2^ or above. Proteinuria was not observed in donor controls, while 32 of 41 (78%) disease controls had spot urine protein-creatinine ratio of 3.0 or above.

**Table 2.** Clinical characteristics of donor and disease controls included in the study. (eGFR: Estimated glomerular filtration rate; N: number, MCD: minimal change disease, MN: membranous nephropathy, FSGS: focal segmental glomerulosclerosis, DN: diabetic nephropathy)

|  | Healthy donor | MCD | MN | FSGS | DN |
| --- | --- | --- | --- | --- | --- |
| number of cases | 7 | 13 | 16 | 6 | 6 |
| age (years) | 56.8 ± 6.2 | 48 ± 12.4 | 52.1 ± 10.3 | 57.2 ± 5.5 | 56 ± 9.8 |
| female (n(%)) | 1 (14.2) | 4 (30.8) | 3 (18.8) | 2 (33.3) | 0 (0) |
| eGFR (mL/min/1.73 m<sup>2</sup>) | 88.9 (88.4-95.5) | 75.3 (48.2-101.3) | 98.6 (85.3-106.6) | 85.0 (80.3-92.8) | 50.7 (40.1-61.7) |
| ≥ 90 (n (%)) | 3 (42.8) | 6 (46.2) | 11 (68.8) | 2 (33.3) | 1 (16.7) |
| ≥ 60 and < 90 (n (%)) | 3 (42.8) | 2 (15.4) | 3 (18.8) | 3 (50.0) | 1 (16.7) |
| ≥ 45 and < 60 (n (%)) | 1 (14.2) | 2 (15.4) | 1 (6.3) | 0 (0) | 1 (16.7) |
| ≥ 30 and < 45 (n (%)) | 0 (0) | 3 (23.1) | 1 (6.3) | 1 (16.7) | 3 (50.0) |
| hemoglobin (g/dL) | 15.0 (14.0-15.3) | 12.8 (12.3-14.3) | 13.0 (11.7-13.5) | 13.4 (11.0-14.7) | 11.3 (10.1-12.5) |
| albumin (mg/dL) | 4.0 (3.8-4.3) | 2 (2-2.4) | 2.5 (1.8-2.8) | 2.4 (2.0-3.6) | 4 (3.5-4.4) |
| ≥ 3.0 (n (%)) | 7 (100) | 1 (7.7) | 3 (18.8) | 4 (66.6) | 5 (83.3) |
| < 3.0 (n (%)) | 0 (0) | 12 (92.3) | 13 (81.2) | 2 (33.3) | 1 (16.7) |
| glucose (mg/dL) | 129 (118-133) | 98 (92-121) | 98 (91-110) | 99 (89-126) | 127 (119-149) |
| spot urine protein-creatinine ratio (g/g) | N/A | 9.3 (6.9-11.3) | 7.4 (4.5-9.0) | 4.5 (1.7-6.0) | 2.7 (1.6-3.1) |
| ≥ 3.0 (n (%)) |  | 12 (92.3) | 14 (87.5) | 4 (66.6) | 2 (33.3) |
| < 3.0 (n (%)) |  | 1 (7.7) | 2 (12.5) | 2 (33.3) | 4 (66.6) |

### Spatial transcriptomics profiling

After RNA-seq processing, 151 glomerular and 51 tubulointerstitial ROIs (3 glomerular ROIs and 1 tubulointerstitial ROIs per specimen, except for one case of MN with 1 glomerular ROI because of the low number of glomeruli available for the profiling) were configured based on histological analysis following hematoxylin and eosin staining and morphological marker staining. In a median (interquartile range) surface area of 31,419 (8,648 to 67,633) and 126,746 (52,430 to 131,871) μm^2^ for each glomerular and tubulointerstitial ROI, respectively, a median of 134 (20 to 250) and 457 (237 to 709) cells/ROI were identified. A median of 104,475 (59,246 to 232,630) reads per ROIs were aligned for RNA-seq and processed for quality control. RNA-seq saturation was 87.9 (interquartile range 81.9, 93.4) %/ROIs. After removal of 58 genes that were expressed in less than 50% of the total samples, 18,677 genes remained for subsequent analyses.

### Differentially expressed genes between C3G and donor control

Differential gene expression analysis was performed between the three C3G samples and the seven healthy donor controls. Compared to the glomeruli of donor controls, 229 genes were upregulated and 563 were downregulated in the glomeruli of C3G samples (**Figure 2A, Supplementary Table 1**), while comparison of tubules between the two groups showed no significant DEGs. Among the upregulated glomerular DEGs, the greatest fold differences were identified in periostin (POSTN) and collagen type I alpha 2 chain (COL1A2). Gene ontology annotations for the upregulated DEGs revealed significant enrichment of multiple molecular function terms including structural molecule activity (GO:0005198), extracellular matrix structural constituent (GO:000520), structural constituent of ribosome (GO:0003735), platelet-derived growth factor binding (GO:0048407), growth factor binding (GO:0019838), and protease binding (GO:0002020) (**Table 3, Supplementary Table 2**). Network analysis of upregulated DEGs with high interaction scores (0.700 threshold) revealed three main clusters: extracellular matrix proteins, ribosomal proteins, and proteins related to interferon activity (**Figure 3A**). Genes KRTCAP3 (keratinocyte associated protein 3), SLC2A14 (solute carrier family 2 member 14), TMEM255A (transmembrane protein 255), DCTPP1 (dCTP pyrophosphatase 1), and SELENOT (Selenoprotein T) were among the significantly downregulated with high fold changes, but no significant functional enrichment could be identified.

**Table 3.**
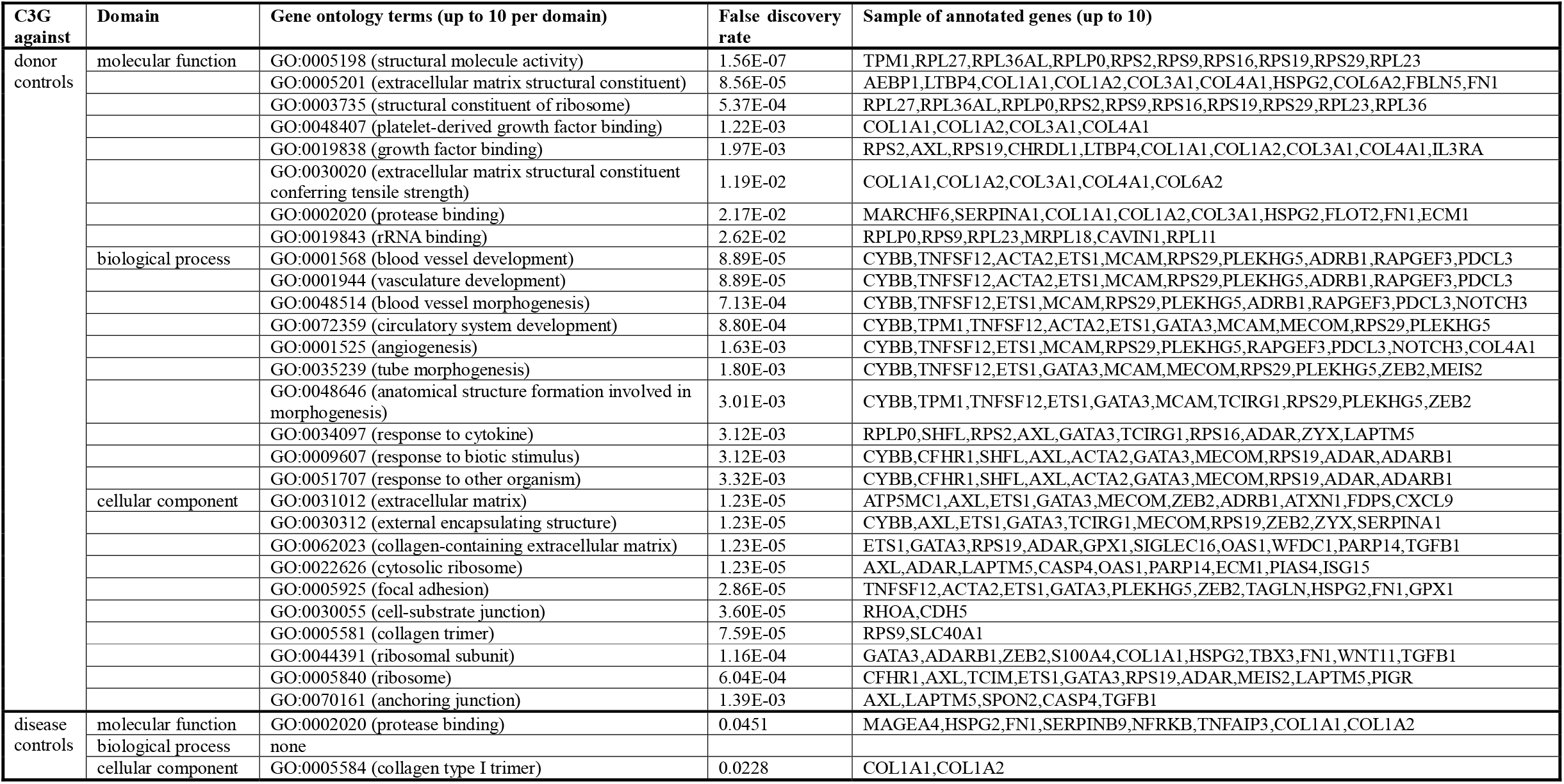
Top gene ontologies among glomerular DEGs upregulated in C3G as annotated by the ToppGene Suite. For each comparison, up to 10 gene ontology terms with the lowest false discovery rate for each domain are presented with a sample of up to 10 individual annotated genes reported by the ToppGene Suite. (C3G: C3 glomerulopathy, DEG: differentially expressed gene)

**Figure 1.**
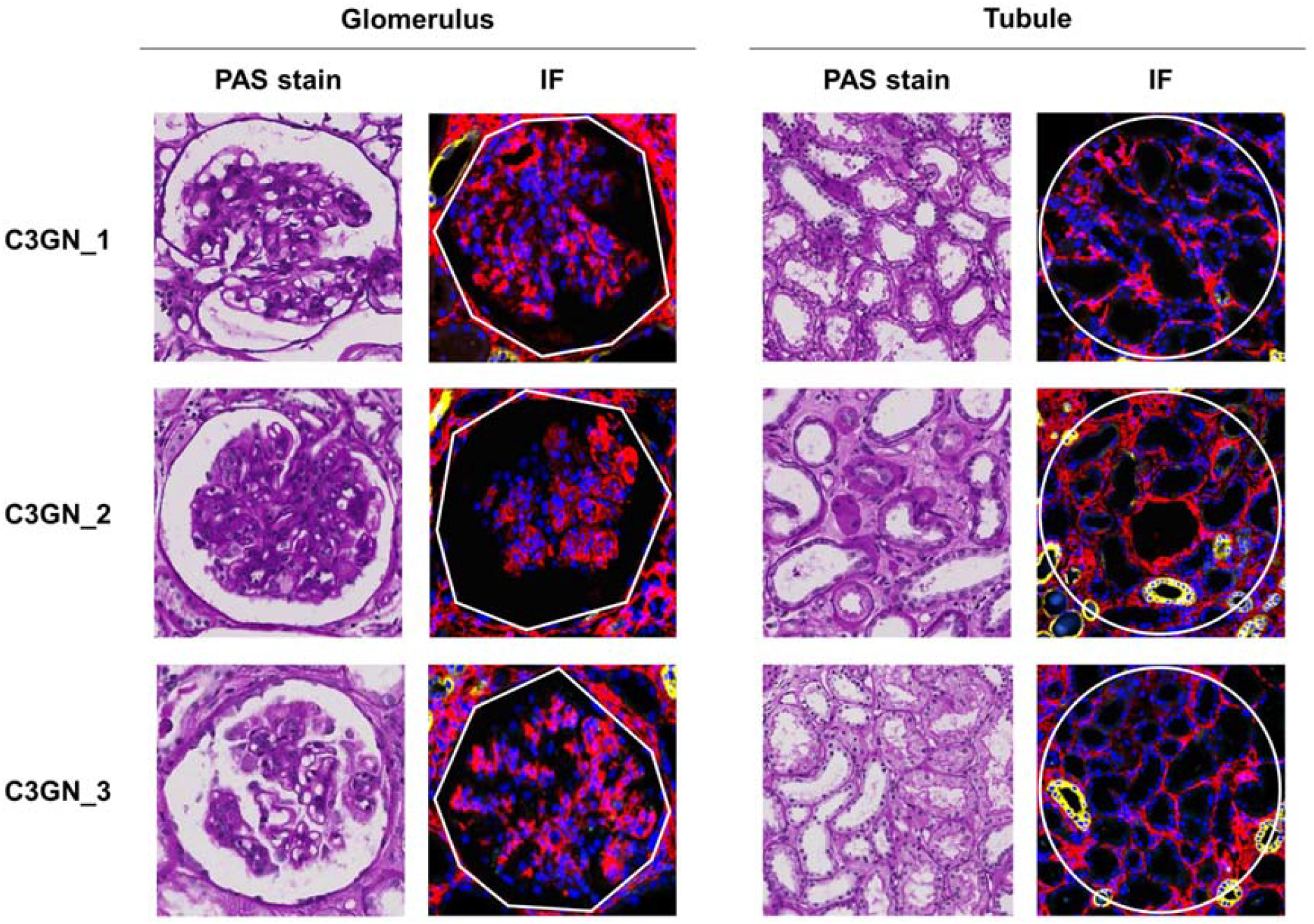
Representative image of the glomerulus and tubule used for spatial transcriptome analysis. PAS-stained image (left) and correspondingly selected ROIs in GeoMx (right). Nuclear (blue), PanCK (yellow) and -SMA (red) were stained to visualize the morphology of each glomerulus. (ROI: region of interest)

**Figure 2.**
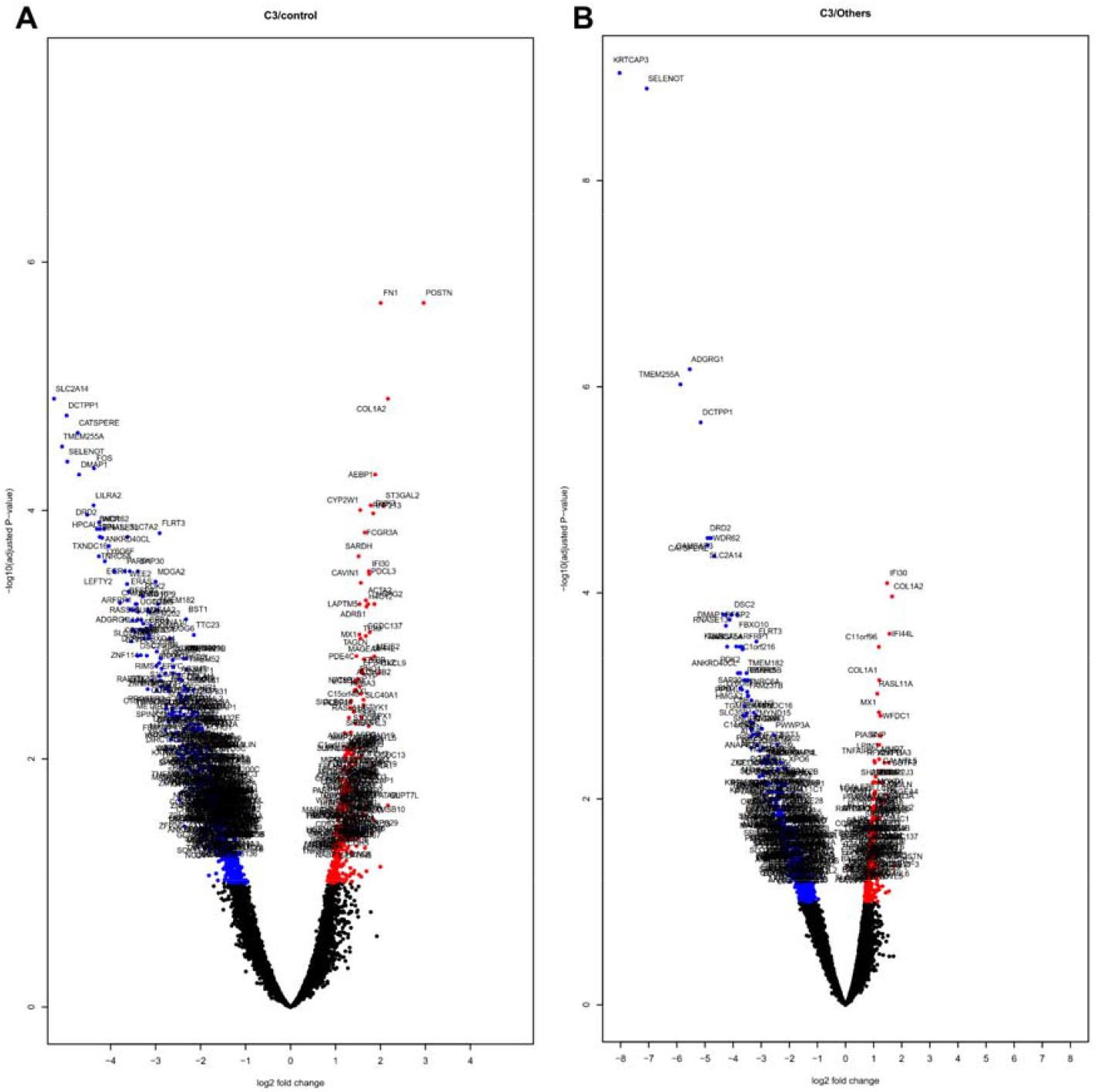
Volcano plots of DEGs between the glomerular transcriptional profile of C3G and the (A) healthy donor controls and (B) other glomerular disease controls. Fold changes in glomerular gene expressions are shown with upregulated and downregulated DEGs with absolute log2 fold change above 1.50 in red and blue, respectively. (DEG: differentially expressed gene)

**Figure 3.**
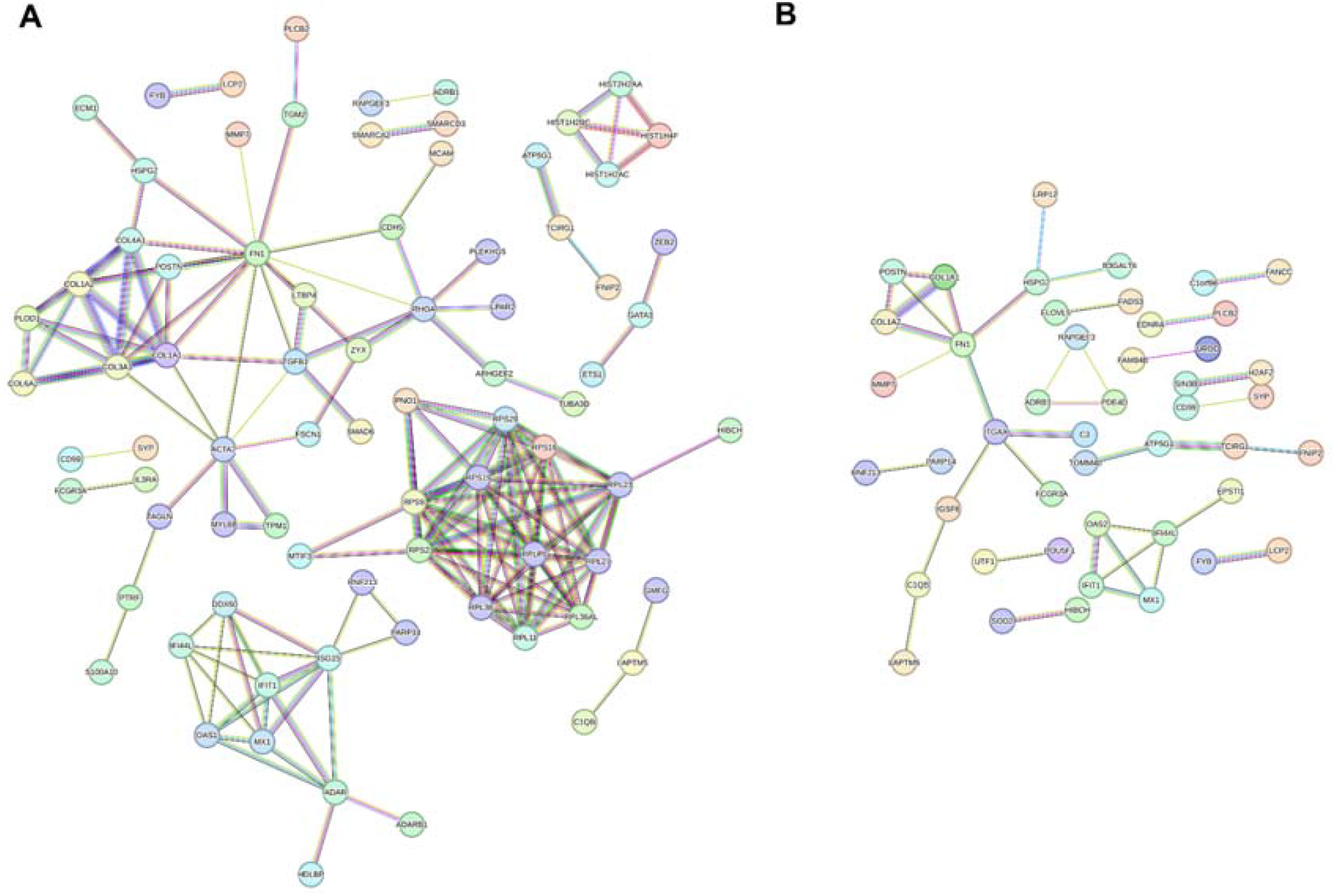
Protein-protein interaction analysis of DEGs in C3G relative to (A) healthy donor controls and (B) other glomerular disease controls. The mapping was performed using the STRING database, and the DEGs with at least one significant interaction of high interaction score (0.700 or above) were displayed in the graph. (DEG: differentially expressed gene)

### Differentially expressed genes between C3G and other major glomerulonephritis cases

Analysis of C3G compared to other major GNs for DEGs showed 157 upregulated and 347 downregulated DEGs in the glomeruli (**Figure 2B, Supplementary Table 1**), and no significant DEGs were found between the tubules. Among the glomerular DEGs, downregulated DEGs showed no significant functional gene enrichment, while upregulated DEGs showed enrichment of the Gene Ontology molecular function term protease binding (GO:0002020) (**Table 3, Supplementary Table 2**). The greatest fold differences among the significantly upregulated DEGs were found in periostin (POSTN), collagen type I alpha 2 chain (COL1A2), cleavage stimulation factor subunit 3 (CSTF3), interferon induced protein 44 like (IFI44L), and regulatory factor X2 (RFX2), of which all but CSTF3 were also upregulated in comparison with the donor controls. Network analysis of upregulated DEGs with high interaction scores (0.700 threshold) showed clusters of extracellular matrix proteins and proteins related to or activated by interferon activity (**Figure 3B**). Notably, C3 occupied a central position in the computed interaction network and showed high probability of interaction with ITGAX (integrin subunit alpha X), which in turn had linkage to the cluster of ECM proteins through high-probability interaction with FN1 and connected to C1QB (complement C1q B chain) through IGSF6 (immunoglobulin superfamily member 6).

## Discussion

In this study, we explored the glomerulus- and tubule-specific transcriptional profile of C3G relative to healthy controls and other glomerular diseases for potential insights into the molecular pathophysiology of C3G. Gene ontology enrichment analysis of the upregulated glomerular DEGs in both comparisons suggested that functional enrichment of genes with protease binding function may be characteristic of C3G. Moreover, clusters of extracellular matrix (ECM) proteins and interferon-related proteins were consistently identified as key components of the protein-protein interaction network among the upregulated DEGs in the glomeruli of C3G. When compared with other glomerular disease controls, the glomerular transcriptional profile C3G had upregulation of complement C3 that had possible interactions with ITGAX, a likewise upregulated integrin subunit protein which in turn had potential interactions with other highly enriched ECM-related proteins in the network, namely FN1, COL1A1, COL1A2, and POSTN.

The observed pattern of DEGs, dominated by upregulation of ECM proteins, appears to be consistent with typical histopathological features of C3G. As disease-specific pathophysiologic mechanisms may mainly occur in glomerulus, there were no prominent tubulointerstitial transcriptomic differences between C3G other glomerulonephritis diseases. In the spatially resolved glomerular transcriptome, the upregulated DEGs with the highest fold changes common to both comparisons were ECM-related proteins POSTN and COL1A2, consistent with the mesangial matrix expansion in the MPGN pattern of injury associated with C3G.^14,15^ While ECM components may also be highly expressed due to the development of glomerular fibrosis in advanced kidney disease, the initial exclusion of samples with low eGFR, manual selection of non-sclerotic glomeruli for spatial analysis, and consistent results found in DEGs relative to other glomerular disease controls altogether make it less likely that their upregulation is a mere consequence of the nonspecific, chronic sclerotic process. In contrast, genes for ribosomal subunits were significantly upregulated in C3G with functional enrichment only when compared to donor controls, suggesting their upregulation may only reflect the increase of protein synthesis due to ongoing glomerular inflammation that is not specific to C3G.

The possible interaction between C3 and ECM proteins suggested by the network analysis could be relevant to the pathophysiology of C3G. Being the namesake of the disease, C3 is expected to show high expression in the glomeruli of C3G,^4^ but the role of the ECM in C3G has not been fully characterized in the existing literature. Dysregulation of the alternative pathway in C3G most frequently occurs in the “fluid phase” of circulating plasma proteins, which subsequently alter the glomerular microenvironment including endothelial cells and the extracellular matrix.^1,4^ Integrin alpha X, encoded by ITGAX, together with CD18 forms complement receptor 4 (CR4), a beta-2 integrin-type complement receptor.^16^ Found on leukocytes, CR4 binds to multiple targets including iC3b and is involved in cellular adhesion, leukocyte migration, and phagocytosis.^17^ Complement receptor 3 (CR3), in the same family of integrins as CR4 and often co-expressed on leukocytes,^18^ has been studied in mouse models of C3G as a potential downregulator of the proinflammatory response through its interactions with iC3b,^19^ and our result suggests that CR4 may also be involved in modulation of inflammation in C3G.

Not typically expressed in normal kidney tissues, periostin is a non-structural component of the ECM that was investigated in multiple previous studies as a potential biomarker of kidney inflammation and fibrosis.^20,21^ As such, the apparent glomerular upregulation of periostin in C3G may indicate not only the ongoing glomerular fibrosis but also the underlying inflammatory process that interacts with the ECM through periostin.

Also among the consistently upregulated DEGs in both comparisons were interferon-related genes with no clear known function in C3G. Namely, the genes IFI44L, IFIT1, MX1, and OAS2 all encode proteins involved in interferon-mediated intracellular antiviral responses.^22^ Activation of type 1 interferon (IFN-I) pathway has been implicated in the development of autoimmune diseases that include lupus nephritis, where IFN-I produced by renal resident cells promote glomerular fibrosis.^23^ A recent analysis identified IFI44, a paralog of IFI44L, as one of the potential key biomarkers of LN among lupus patients.^24^Although IFN-I activation has not been previously associated with C3G, the findings suggest that interferon-related signals may have pro-inflammatory role in C3G similar to LN, contributing to the glomerular pathology of the disease.

There are several limitations in this study. First, as our study was based on three selected cases of C3G in male Korean adults, selection bias may exist. As C3G shows diverse clinical courses, additional effort is warranted to reveal the high-risk clinicopathologic characteristics that may be used to stratify the disease category. Second, protein-level validation experiment was not performed, as the main translated proteins from the identified DEGs are mostly difficult to be quantified in the glomerulus. On the other hand, this may suggest that the current spatial transcriptomic assay was able to identify early mRNA-level pathologic changes in the glomerulus of C3G. Third, single-cell level interpretation of the result was limited. This was because, as C3G is a rare disease, performing single-cell RNA sequencing is difficult due to the limited availability of a fresh kidney biopsy sample. In addition, most single-cell level data of kidney biopsy tissue includes a limited number of glomerular cells; thus, single-cell RNA sequencing may not be an ideal method to investigate intraglomerular transcriptomic alteration of such rare disease as C3G.

In conclusion, we explored the spatial transcriptomic profile of C3G and identified, among differentially expressed genes in the glomerulus, consistent enrichment of genes related to the extracellular matrix and to interferon activity. As the first report of kidney substructure-specific transcriptomic profile of C3GN to date, our study suggests these upregulated genes may have a previously underrecognized role in the complex complement-mediated pathogenesis of C3G.

## Data Availability

All data produced in the present study are available upon reasonable request to the authors.

## Acknowledgements

This work was supported by the National Research Foundation of Korea (NRF) grant funded by the Korea government (MSIT, Ministry of Science and ICT) (No. 2021R1A2C2094586) and by the Bio & Medical Technology Development Program of the National Research Foundation (NRF) funded by the Korean government (MSIT) (2022M3A9D3016848).

## Supplemental Contents

Supplementary Table 1. All DEGs between C3G cases and healthy donor controls and between C3G cases and other glomerulonephritis controls by DESeq2 method. Log2 fold changes and FDR are presented for each significant DEG. (DEG: differentially expressed genes, C3G: C3 glomerulopathy, FDR: false discovery rate)

Supplementary Table 2. Significant gene ontologies among upregulated DEGs as annotated by the ToppGene Suite. For each comparison, all gene ontology terms with the lowest false discovery rate for each domain are presented with all individual annotated genes reported by the ToppGene Suite. (C3G: C3 glomerulopathy)

