## Supplementary Table 2 for "Spatially resolved transcriptomic profiling for glomerular and tubulointerstitial gene expression in C3 glomerulopathy"

Supplementary Table 2. Significant gene ontologies among upregulated DEGs as annotated by the ToppGene Suite. For each comparison, all gene ontology terms with the lowest false discovery rate for each domain are presented with all individual annotated genes reported by the ToppGene Suite. (C3G: C3 glomerulopathy)

| C3G against | Domain | Gene ontology terms | False discovery rate | Annotated gene lists |
| --- | --- | --- | --- | --- |
| donor controls | molecular function | GO:0005198 (structural molecule activity) | 1.56E-07 | TPM1,RPL27,RPL36AL,RPLP0,RPS2,RPS9,RPS16,RPS19,RPS29,RPL23,H2AC6,H2BC4,AEBP1,H4C12,MYL6B,LTBP4,COL1A1,COL1A2,COL3A1,COL4A1,HSPG2,COL6A2,RPL36,FBLN5,FN1,SEPTIN5,TUBA3D,H2AC19,ECM1,POSTN,MRPL18,MMRN2,CAVIN1,EPB41L2,RPL11 |
|  |  | GO:0005201 (extracellular matrix structural constituent) | 8.56E-05 | AEBP1,LTBP4,COL1A1,COL1A2,COL3A1,COL4A1,HSPG2,COL6A2,FBLN5,FN1,ECM1,POSTN,MMRN2 |
|  |  | GO:0003735 (structural constituent of ribosome) | 5.37E-04 | RPL27,RPL36AL,RPLP0,RPS2,RPS9,RPS16,RPS19,RPS29,RPL23,RPL36,MRPL18,RPL11 |
|  |  | GO:0048407 (platelet-derived growth factor binding) | 1.22E-03 | COL1A1,COL1A2,COL3A1,COL4A1 |
|  |  | GO:0019838 (growth factor binding) | 1.97E-03 | RPS2,AXL,RPS19,CHRDL1,LTBP4,COL1A1,COL1A2,COL3A1,COL4A1,IL3RA |
|  |  | GO:0030020 (extracellular matrix structural constituent conferring tensile strength) | 1.19E-02 | COL1A1,COL1A2,COL3A1,COL4A1,COL6A2 |
|  |  | GO:0002020 (protease binding) | 2.17E-02 | MARCHF6,SERPINA1,COL1A1,COL1A2,COL3A1,HSPG2,FLOT2,FN1,ECM1 |
|  |  | GO:0019843 (rRNA binding) | 2.62E-02 | RPLP0,RPS9,RPL23,MRPL18,CAVIN1,RPL11 |
|  | biological process | GO:0001568 (blood vessel development) | 8.89E-05 | CYBB,TNFSF12,ACTA2,ETS1,MCAM,RPS29,PLEKHG5,ADRB1,RAPGEF3,PDCL3,NOTCH3,COL1A1,COL1A2,COL3A1,COL4A1,HSPG2,TBX3,FBLN5,FN1,WNT11,GPX1,GMFG,SOX17,RNF213,NIBAN2,ECM1,TGFB1,RHOA,PPP3R1,MMRN2,LPAR2,TNFAIP2,PEAK1,CDH5,GAA,SMAD6 |
|  |  | GO:0001944 (vasculature development) | 8.89E-05 | CYBB,TNFSF12,ACTA2,ETS1,MCAM,RPS29,PLEKHG5,ADRB1,RAPGEF3,PDCL3,NOTCH3,COL1A1,COL1A2,COL3A1,COL4A1,HSPG2,TBX3,FBLN5,FN1,WNT11,GPX1,GMFG,SOX17,RNF213,NIBAN2,ECM1,TGFB1,RHOA,PPP3R1,SMARCA2,MMRN2,LPAR2,TNFAIP2,PEAK1,CDH5,GAA,SMAD6 |
|  |  | GO:0048514 (blood vessel morphogenesis) | 7.13E-04 | CYBB,TNFSF12,ETS1,MCAM,RPS29,PLEKHG5,ADRB1,RAPGEF3,PDCL3,NOTCH3,COL3A1,COL4A1,HSPG2,FBLN5,FN1,WNT11,GPX1,GMFG,SOX17,RNF213,NIBAN2,ECM1,TGFB1,RHOA,PPP3R1,MMRN2,LPAR2,TNFAIP2,PEAK1,CDH5,SMAD6 |
|  |  | GO:0072359 (circulatory system development) | 8.80E-04 | CYBB,TPM1,TNFSF12,ACTA2,ETS1,GATA3,MCAM,MECOM,RPS29,PLEKHG5,MEIS2,ADRB1,RAPGEF3,FDPS,PDCL3,PLCB2,NOTCH3,COL1A1,COL1A2,COL3A1,COL4A1,HSPG2,TBX3,FBLN5,FN1,WNT11,GPX1,GMFG,SOX17,RNF213,NIBAN2,ECM1,CRIP1,TGFB1,RHOA,PPP3R1,WDR11,SMARCA2,MMRN2,SMARCD3,LPAR2,TNFAIP2,PEAK1,CDH5,GAA,SMAD6 |
|  |  | GO:0001525 (angiogenesis) | 1.63E-03 | CYBB,TNFSF12,ETS1,MCAM,RPS29,PLEKHG5,RAPGEF3,PDCL3,NOTCH3,COL4A1,HSPG2,FBLN5,FN1,GPX1,GMFG,SOX17,RNF213,NIBAN2,ECM1,TGFB1,RHOA,PPP3R1,MMRN2,LPAR2,TNFAIP2,PEAK1,CDH5 |
|  |  | GO:0035239 (tube morphogenesis) | 1.80E-03 | CYBB,TNFSF12,ETS1,GATA3,MCAM,MECOM,RPS29,PLEKHG5,ZEB2,MEIS2,ADRB1,RAPGEF3,PDCL3,NOTCH3,COL3A1,COL4A1,HSPG2,TBX3,FBLN5,FN1,WNT11,GPX1,GMFG,SOX17,RNF213,NIBAN2,ECM1,TGFB1,RHOA,TGM2,PPP3R1,MMRN2,LPAR2,TNFAIP2,PEAK1,CDH5,SMAD6 |
|  |  | GO:0048646 (anatomical structure formation involved in morphogenesis) | 3.01E-03 | CYBB,TPM1,TNFSF12,ETS1,GATA3,MCAM,TCIRG1,RPS29,PLEKHG5,ZEB2,RAPGEF3,PDCL3,CXCL9,NOTCH3,COL1A1,COL3A1,COL4A1,HSPG2,TBX3,FBLN5,FN1,WNT11,GPX1,GMFG,SOX17,RNF213,NPC2,NIBAN2,ECM1,RFX2,SLC40A1,TGFB1,RHOA,TGM2,PPP3R1,MMRN2,SMARCD3,LPAR2,TNFAIP2,PEAK1,PLEKHO1,CDH5,PRRC1 |
|  |  | GO:0034097 (response to cytokine) | 3.12E-03 | RPLP0,SHFL,RPS2,AXL,GATA3,TCIRG1,RPS16,ADAR,ZYX,LAPTM5,SERPINA1,CXCL9,COL1A1,COL3A1,FN1,SOX17,CASP4,OAS1,PARP14,ECM1,RFX2,IFIT1,RHOA,PIAS4,ISG15,LSP1,ARHGEF2,IL3RA,MX1,HDGF |
|  |  | GO:0009607 (response to biotic stimulus) | 3.12E-03 | CYBB,CFHR1,SHFL,AXL,ACTA2,GATA3,MECOM,RPS19,ADAR,ADARB1,ZYX,LAPTM5,H2AC6,H2BC4,FCGR3A,SPON2,CXCL9,C1QB,IFI44L,MMP7,FLOT2,WIPF1,DDX60,GPX1,SIGLEC16,H2AC19,CASP4,RNF213,OAS1,NPC2,PARP14,IFIT1,TGFB1,RHOA,PIAS4,ISG15,C15orf48,MMRN2,TMSB10,INAVA,ARHGEF2,MX1,RPL11,SMAD6 |
|  |  | GO:0051707 (response to other organism) | 3.32E-03 | CYBB,CFHR1,SHFL,AXL,ACTA2,GATA3,MECOM,RPS19,ADAR,ADARB1,ZYX,H2AC6,H2BC4,FCGR3A,SPON2,CXCL9,C1QB,IFI44L,MMP7,FLOT2,WIPF1,DDX60,GPX1,SIGLEC16,H2AC19,CASP4,RNF213,OAS1,NPC2,PARP14,IFIT1,TGFB1,RHOA,PIAS4,ISG15,C15orf48,MMRN2,TMSB10,INAVA,ARHGEF2,MX1,RPL11,SMAD6 |
|  |  | GO:0043207 (response to external biotic stimulus) | 3.32E-03 | CYBB,CFHR1,SHFL,AXL,ACTA2,GATA3,MECOM,RPS19,ADAR,ADARB1,ZYX,H2AC6,H2BC4,FCGR3A,SPON2,CXCL9,C1QB,IFI44L,MMP7,FLOT2,WIPF1,DDX60,GPX1,SIGLEC16,H2AC19,CASP4,RNF213,OAS1,NPC2,PARP14,IFIT1,TGFB1,RHOA,PIAS4,ISG15,C15orf48,MMRN2,TMSB10,INAVA,ARHGEF2,MX1,RPL11,SMAD6 |
|  |  | GO:0098542 (defense response to other organism) | 3.32E-03 | CYBB,CFHR1,SHFL,AXL,GATA3,RPS19,ADAR,ADARB1,ZYX,H2AC6,H2BC4,FCGR3A,SPON2,CXCL9,C1QB,IFI44L,MMP7,FLOT2,DDX60,SIGLEC16,H2AC19,CASP4,RNF213,OAS1,PARP14,IFIT1,TGFB1,PIAS4,ISG15,MMRN2,TMSB10,INAVA,ARHGEF2,MX1,RPL11 |
|  |  | GO:0035295 (tube development) | 4.57E-03 | CYBB,TNFSF12,ETS1,GATA3,MCAM,MECOM,RPS29,PLEKHG5,ZEB2,MEIS2,ADRB1,ATXN1,RAPGEF3,PDCL3,FKBP8,NOTCH3,COL3A1,COL4A1,HSPG2,TBX3,FBLN5,FN1,WNT11,GPX1,GMFG,SOX17,RNF213,NPC2,NIBAN2,ECM1,TGFB1,RHOA,TGM2,PPP3R1,MMRN2,LPAR2,TNFAIP2,PEAK1,FSCN1,CDH5,SMAD6 |
|  |  | GO:0035455 (response to interferon-alpha) | 4.57E-03 | AXL,GATA3,ADAR,OAS1,IFIT1 |
|  |  | GO:0071345 (cellular response to cytokine stimulus) | 5.18E-03 | RPLP0,RPS2,AXL,GATA3,TCIRG1,RPS16,ADAR,ZYX,LAPTM5,CXCL9,COL1A1,FN1,SOX17,CASP4,OAS1,PARP14,ECM1,RFX2,IFIT1,RHOA,PIAS4,ISG15,LSP1,ARHGEF2,IL3RA,MX1,HDGF |
|  |  | GO:0002181 (cytoplasmic translation) | 6.23E-03 | RPL27,RPLP0,RPS2,RPS9,RPS16,RPS19,RPS29,RPL23,RPL36,RPL11 |
|  |  | GO:0140236 (translation at presynapse) | 6.31E-03 | RPL27,RPLP0,RPS16,RPL23,RPL36,RPL11 |
|  |  | GO:0140241 (translation at synapse) | 6.31E-03 | RPL27,RPLP0,RPS16,RPL23,RPL36,RPL11 |
|  |  | GO:0140242 (translation at postsynapse) | 6.31E-03 | RPL27,RPLP0,RPS16,RPL23,RPL36,RPL11 |
|  |  | GO:0035457 (cellular response to interferon-alpha) | 6.55E-03 | AXL,GATA3,OAS1,IFIT1 |
|  |  | GO:0048871 (multicellular organismal-level homeostasis) | 7.84E-03 | RPL27,AXL,RPS9,ETS1,GATA3,TCIRG1,MECOM,RPS19,RPS29,ADAR,TUB,ZEB2,ADRB1,PIGR,FCGR3A,TPP1,NOTCH3,COL3A1,GPX1,OAS1,SLC40A1,TGFB1,ISG15,SMARCA2,LPAR2,INAVA,CDH5,PRRC1,RPL11 |
|  |  | GO:2000026 (regulation of multicellular organismal development) | 7.84E-03 | CYBB,ATP5MC1,BTBD6,TNFSF12,AXL,TCIM,ETS1,GATA3,RPS19,ZEB2,MEIS2,S100A10,ADRB1,ATXN1,H4C12,RAPGEF3,FDPS,PDCL3,NOTCH3,HSPG2,TBX3,FBLN5,FN1,WNT11,SOX17,RNF213,NIBAN2,ECM1,TGFB1,RHOA,TGM2,NSMF,ISG15,TM4SF4,SMARCA2,MMRN2,SMARCD3,LPAR2,MAP3K13,ARHGEF2,CDH5 |
|  |  | GO:0034101 (erythrocyte homeostasis) | 7.91E-03 | RPL27,AXL,RPS9,ETS1,GATA3,RPS19,RPS29,ADAR,SLC40A1,ISG15,LPAR2,RPL11 |
|  |  | GO:0016049 (cell growth) | 8.09E-03 | ATP5MC1,ZEB2,S100A10,ADRB1,FDPS,LTBP4,FBLN5,FN1,SYT3,WNT11,SOX17,WFDC1,SIPA1,TGFB1,RHOA,POSTN,NSMF,SMARCA2,MAP3K13,IL3RA,CDH5 |
|  |  | GO:0043542 (endothelial cell migration) | 8.61E-03 | TNFSF12,ETS1,GATA3,PLEKHG5,ZEB2,TAGLN,HSPG2,FN1,GPX1,TGFB1,RHOA,MMRN2,PEAK1,CDH5 |
|  |  | GO:0045087 (innate immune response) | 8.61E-03 | CYBB,SHFL,AXL,GATA3,RPS19,ADAR,ADARB1,ZYX,H2AC6,H2BC4,FCGR3A,SPON2,C1QB,FLOT2,DDX60,SIGLEC16,H2AC19,CASP4,OAS1,PARP14,IFIT1,TGFB1,PIAS4,ISG15,INAVA,ARHGEF2,MX1,RPL11 |
|  |  | GO:0043009 (chordate embryonic development) | 8.74E-03 | TPM1,RPL36AL,RPLP0,GATA3,MECOM,RPS19,RPS29,ADAR,ADARB1,ZEB2,MEIS2,SERPINA1,SLC35E2A,FKBP8,NOTCH3,COL1A1,COL3A1,HSPG2,TBX3,NAT8B,FN1,WNT11,SOX17,CASP4,APBA3,TGFB1,CDK2AP1,FSCN1,PRRC1,RPL11 |
|  |  | GO:0097435 (supramolecular fiber organization) | 8.74E-03 | TPM1,ZEB2,ZYX,TBCB,S100A10,CCDC13,AEBP1,RAPGEF3,TAGLN,LTBP4,COL1A1,COL1A2,COL3A1,HSPG2,NAT8B,FBLN5,WIPF1,WNT11,GPX1,GMFG,CASP4,RHOA,TMSB10,ARHGEF2,FSCN1,CDH5 |
|  |  | GO:0045071 (negative regulation of viral genome replication) | 9.14E-03 | SHFL,ADAR,OAS1,IFIT1,ISG15,MX1 |
|  |  | GO:0030010 (establishment of cell polarity) | 9.47E-03 | GATA3,TCIRG1,FLOT2,FN1,WNT11,SIPA1,RHOA,ARHGEF2,FSCN1,CDH5 |
|  |  | GO:0001558 (regulation of cell growth) | 9.47E-03 | ATP5MC1,S100A10,ADRB1,FDPS,LTBP4,FBLN5,FN1,SYT3,WNT11,SOX17,WFDC1,SIPA1,TGFB1,RHOA,NSMF,SMARCA2,MAP3K13,IL3RA |
|  |  | GO:0009792 (embryo development ending in birth or egg hatching) | 1.03E-02 | TPM1,RPL36AL,RPLP0,GATA3,MECOM,RPS19,RPS29,ADAR,ADARB1,ZEB2,MEIS2,SERPINA1,SLC35E2A,FKBP8,NOTCH3,COL1A1,COL3A1,HSPG2,TBX3,NAT8B,FN1,WNT11,SOX17,CASP4,APBA3,TGFB1,CDK2AP1,FSCN1,PRRC1,RPL11 |
|  |  | GO:0002262 (myeloid cell homeostasis) | 1.03E-02 | RPL27,AXL,RPS9,ETS1,GATA3,MECOM,RPS19,RPS29,ADAR,SLC40A1,ISG15,LPAR2,RPL11 |
|  |  | GO:0034340 (response to type I interferon) | 1.10E-02 | SHFL,ADAR,OAS1,IFIT1,PIAS4,ISG15,MX1 |
|  |  | GO:0060339 (negative regulation of type I interferon-mediated signaling pathway) | 1.10E-02 | ADAR,OAS1,PIAS4,ISG15 |
|  |  | GO:0045069 (regulation of viral genome replication) | 1.27E-02 | SHFL,ADAR,ADARB1,OAS1,IFIT1,ISG15,MX1 |
|  |  | GO:0071229 (cellular response to acid chemical) | 1.36E-02 | CYBB,ATP5MC1,S100A10,COL1A1,COL1A2,COL3A1,COL4A1,SIPA1,NSMF |
|  |  | GO:0030218 (erythrocyte differentiation) | 1.38E-02 | RPL27,RPS9,ETS1,GATA3,RPS19,RPS29,ADAR,SLC40A1,ISG15,LPAR2,RPL11 |
|  |  | GO:0030198 (extracellular matrix organization) | 1.44E-02 | ETS1,AEBP1,MMP7,LTBP4,COL1A1,COL1A2,COL3A1,COL4A1,HSPG2,FBLN5,FN1,TGFB1,POSTN,FSCN1 |
|  |  | GO:0043062 (extracellular structure organization) | 1.44E-02 | ETS1,AEBP1,MMP7,LTBP4,COL1A1,COL1A2,COL3A1,COL4A1,HSPG2,FBLN5,FN1,TGFB1,POSTN,FSCN1 |
|  |  | GO:0045229 (external encapsulating structure organization) | 1.52E-02 | ETS1,AEBP1,MMP7,LTBP4,COL1A1,COL1A2,COL3A1,COL4A1,HSPG2,FBLN5,FN1,TGFB1,POSTN,FSCN1 |
|  |  | GO:0040007 (growth) | 1.61E-02 | ATP5MC1,AXL,GATA3,ADARB1,ZEB2,S100A10,ADRB1,FDPS,FKBP8,LTBP4,COL1A1,COL3A1,NAT8B,FBLN5,FN1,SYT3,WNT11,GPX1,SOX17,WFDC1,SIPA1,ECM1,TGFB1,RHOA,POSTN,NSMF,WDR11,TM4SF4,SMARCA2,MAP3K13,IL3RA,CDH5 |
|  |  | GO:0001101 (response to acid chemical) | 1.93E-02 | CYBB,ATP5MC1,S100A10,COL1A1,COL1A2,COL3A1,COL4A1,SIPA1,RHOA,NSMF |
|  |  | GO:0043534 (blood vessel endothelial cell migration) | 2.03E-02 | ETS1,PLEKHG5,HSPG2,FN1,GPX1,TGFB1,RHOA,MMRN2,PEAK1,CDH5 |
|  |  | GO:0010935 (regulation of macrophage cytokine production) | 2.06E-02 | AXL,LAPTM5,SPON2,CASP4,TGFB1 |
|  |  | GO:0001959 (regulation of cytokine-mediated signaling pathway) | 2.06E-02 | AXL,ADAR,LAPTM5,CASP4,OAS1,PARP14,ECM1,PIAS4,ISG15 |
|  |  | GO:0009617 (response to bacterium) | 2.10E-02 | AXL,MECOM,RPS19,H2AC6,H2BC4,FCGR3A,SPON2,CXCL9,MMP7,GPX1,SIGLEC16,H2AC19,CASP4,RNF213,OAS1,IFIT1,TGFB1,RHOA,ISG15,C15orf48,MMRN2,TMSB10,INAVA,MX1,SMAD6 |
|  |  | GO:0085029 (extracellular matrix assembly) | 2.35E-02 | LTBP4,COL1A2,COL3A1,FBLN5,TGFB1 |
|  |  | GO:0048251 (elastic fiber assembly) | 2.35E-02 | LTBP4,COL3A1,FBLN5 |
|  |  | GO:0060284 (regulation of cell development) | 2.35E-02 | ATP5MC1,BTBD6,AXL,TCIM,ETS1,GATA3,RPS19,ZEB2,MEIS2,S100A10,ATXN1,H4C12,NOTCH3,FN1,NIBAN2,TGFB1,RHOA,TGM2,NSMF,ISG15,TM4SF4,SMARCA2,SMARCD3,LPAR2,MAP3K13,ARHGEF2,CDH5 |
|  | cellular component | GO:0031012 (extracellular matrix) | 1.23E-05 | ATP5MC1,AXL,ETS1,GATA3,MECOM,ZEB2,ADRB1,ATXN1,FDPS,CXCL9,ADIRF,COL1A1,FN1,SOX17,NIBAN2,TGFB1,MSR1,RHOA,SLC6A6,TGM2,NSMF,ISG15,SMARCA2,SMARCD3,MAP3K13,ARHGEF2,CDH5 |
|  |  | GO:0030312 (external encapsulating structure) | 1.23E-05 | CYBB,AXL,ETS1,GATA3,TCIRG1,MECOM,RPS19,ZEB2,ZYX,SERPINA1,FCGR3A,CXCL9,HSPG2,FN1,GPX1,GMFG,SIGLEC16,CASP4,WFDC1,NPC2,ECM1,TGFB1,TGM2,TMSB10,CDH5 |
|  |  | GO:0062023 (collagen-containing extracellular matrix) | 1.23E-05 | ETS1,GATA3,RPS19,ADAR,GPX1,SIGLEC16,OAS1,WFDC1,PARP14,TGFB1,PIAS4,ISG15,CDH5 |
|  |  | GO:0022626 (cytosolic ribosome) | 1.23E-05 | AXL,ADAR,LAPTM5,CASP4,OAS1,PARP14,ECM1,PIAS4,ISG15 |
|  |  | GO:0005925 (focal adhesion) | 2.86E-05 | TNFSF12,ACTA2,ETS1,GATA3,PLEKHG5,ZEB2,TAGLN,HSPG2,FN1,GPX1,TGFB1,RHOA,MMRN2,PEAK1,CDH5 |
|  |  | GO:0030055 (cell-substrate junction) | 3.60E-05 | RHOA,CDH5 |
|  |  | GO:0005581 (collagen trimer) | 7.59E-05 | RPS9,SLC40A1 |
|  |  | GO:0044391 (ribosomal subunit) | 1.16E-04 | GATA3,ADARB1,ZEB2,S100A4,COL1A1,HSPG2,TBX3,FN1,WNT11,TGFB1,PPP3R1,WDR11,FSCN1 |
|  |  | GO:0005840 (ribosome) | 6.04E-04 | CFHR1,AXL,TCIM,ETS1,GATA3,RPS19,ADAR,MEIS2,LAPTM5,PIGR,FCGR3A,H4C12,CD99,SPON2,C1QB,COL3A1,FLOT2,FN1,DDX60,GPX1,SIGLEC16,CASP4,OAS1,PARP14,LCP2,ECM1,TGFB1,RHOA,PIAS4,ISG15,SMARCA2,SMARCD3,LPAR2,INAVA,FYB1,RPL11 |
|  |  | GO:0070161 (anchoring junction) | 1.39E-03 | AXL,LAPTM5,SPON2,CASP4,TGFB1 |
|  |  | GO:0022625 (cytosolic large ribosomal subunit) | 1.83E-03 | TNFSF12,ACTA2,ETS1,GATA3,PLEKHG5,ZEB2,TAGLN,HSPG2,FN1,WNT11,GPX1,SOX17,TGFB1,RHOA,WDR11,MMRN2,PEAK1,FSCN1,CDH5 |
|  |  | GO:0015629 (actin cytoskeleton) | 1.97E-03 | RPL27,AXL,RPS9,ETS1,GATA3,TCIRG1,MECOM,RPS19,RPS29,ADAR,SLC40A1,TGFB1,ISG15,SMARCA2,LPAR2,RPL11 |
|  |  | GO:0098644 (complex of collagen trimers) | 2.52E-03 | RPS19,ADAR,OAS1,PARP14,TGFB1,PIAS4,ISG15 |
|  |  | GO:0098556 (cytoplasmic side of rough endoplasmic reticulum membrane) | 2.83E-03 | ACTA2,GATA3,ADARB1,ZEB2,S100A4,NOTCH3,COL1A1,HSPG2,TBX3,FN1,WNT11,TGFB1,PPP3R1,WDR11,FSCN1 |
|  |  | GO:0005584 (collagen type I trimer) | 3.63E-03 | CYBB,ATP5MC1,TNFSF12,AXL,ETS1,GATA3,MECOM,ZEB2,ADRB1,ATXN1,RAPGEF3,FDPS,PDCL3,CXCL9,ADIRF,COL1A1,FN1,SYT3,WNT11,SOX17,NIBAN2,ECM1,TGFB1,MSR1,RHOA,SLC6A6,TGM2,NSMF,ISG15,SMARCA2,MMRN2,SMARCD3,MAP3K13,ARHGEF2,CDH5 |
|  |  | GO:0030867 (rough endoplasmic reticulum membrane) | 3.63E-03 | ADAR,OAS1,IFIT1,PIAS4,ISG15,MX1 |
|  |  | GO:0022627 (cytosolic small ribosomal subunit) | 5.67E-03 | TPM1,S100A10,RAPGEF3,WNT11,RHOA |
|  |  | GO:0005583 (fibrillar collagen trimer) | 6.33E-03 | CXCL9,TBX3,FLOT2,TGFB1,SMARCA2,SMARCD3 |
|  |  | GO:0098643 (banded collagen fibril) | 6.33E-03 | TCIRG1,ADAR,H4C12,MRC2,CHRDL1,COL1A1,COL1A2,HSPG2,FN1,WNT11,ECM1,SLC40A1,TGFB1,RHOA,ISG15,SMAD6 |
|  |  | GO:0015934 (large ribosomal subunit) | 7.65E-03 | ADAR,MEIS2,CHRDL1,TBX3,FN1,PRRC1,SMAD6 |
|  |  | GO:0098554 (cytoplasmic side of endoplasmic reticulum membrane) | 9.62E-03 | CFHR1,GATA3,RPS19,ADAR,LAPTM5,PIGR,FCGR3A,C1QB,COL3A1,FLOT2,DDX60,GPX1,SIGLEC16,CASP4,OAS1,PARP14,LCP2,ECM1,TGFB1,PIAS4,ISG15,INAVA,FYB1,RPL11 |
|  |  | GO:0042788 (polysomal ribosome) | 1.16E-02 | TPM1,ZEB2,ZYX,S100A10,RAPGEF3,WNT11,RHOA |
|  |  | GO:0043202 (lysosomal lumen) | 2.17E-02 | TPM1,ZEB2,ZYX,S100A10,RAPGEF3,WNT11,RHOA |
|  |  | GO:0098794 (postsynapse) | 2.81E-02 | SHFL,AXL,ADAR,ADARB1,OAS1,IFIT1,ISG15,MX1 |
|  |  | GO:0015935 (small ribosomal subunit) | 4.61E-02 | ADAR,OAS1,PARP14,ECM1,PIAS4,ISG15 |
| disease controls | molecular function | GO:0002020 (protease binding) | 0.0451 | MAGEA4,HSPG2,FN1,SERPINB9,NFRKB,TNFAIP3,COL1A1,COL1A2 |
|  | biological process | N/A |  |  |
|  | cellular component | GO:0005584 (collagen type I trimer) | 0.0228 | COL1A1,COL1A2 |
